# A study protocol for an interrupted time series analysis and pre-post surveys to assess the effects a community-wide sanitation system on environmental contamination, infection risk, and well-being in Alabama’s Black Belt

**DOI:** 10.1101/2025.07.25.25332164

**Authors:** Olivia A. Harmon, Megan E.J. Lott, David A. Holcomb, Emily McGlohn, Mark Elliott, Kevin White, Joe Brown

## Abstract

**Introduction:** Rural sanitation deficits in the United States represent an important source of non-point source pollution and may present risks to public health. We propose an interrupted time series analysis to measure the effect of a town-wide sanitation expansion program on the release of pathogens to the environment. This work is expected to yield valuable insight into the potential for rural sanitation improvements to reduce pathogen releases and support public health and well-being.

**Methods:** We will conduct a longitudinal baseline study including quantitative measurement of key enteric pathogens and fecal indicator bacteria adjacent to households lacking adequate sanitation. As households connect to a new sewerage system serving the entire community, longitudinal household sampling will continue until crossover is complete. We will include concurrent comparison sites with existing appropriate sanitation as well as sites never receiving the intervention to monitor secular trends in pathogen releases during the study period.

**Analysis:** We will compare the concentration of culturable *E. coli* in the environment pre- and post-intervention using a time series regression analysis suitable for an interrupted time series. We will couple pathogen measurements with quantitative microbial risk assessment to estimate the potential effect of the intervention on infection risks via key exposure pathways. A linked pre-post survey will focus on self-reported quality of life measures among households connecting to the system.

**Ethics and dissemination:** Informed consent will be obtained prior to data collection, with participants informed of study details and risks. Participation is completely voluntary, and identifiable data will be securely and separately stored from all other data. Each household will be offered a summary of their site-specific data. Deidentified results will be shared with the community in a public forum and published in peer-reviewed journals.

**Strengths and Limitations of this Study:**

- This study utilizes both molecular and culture-based environmental sampling with structured household and observational surveys to carefully assess the effects of a community-wide sanitation system on environmental contamination, infection risk, and well-being.
- To address potential confounding due to secular trends and/or weather-related events, we plan to include at least 10 comparison sites with existing improved sanitation
- infrastructure and 10 sites with poor sanitation infrastructure not receiving the intervention during the study period.
- Bias will be reduced by defining enteric pathogen targets and the main statistical models prior to analysis.
- Although we will adjust for external trends such as temperature and rainfall, the design of an interrupted time series lacks a randomized control group, limiting the ability to fully isolate the intervention effect.
- Voluntary participation and required utility fees may lead to selection bias, and survey responses may be influenced by social desirability or courtesy bias. We aim to mitigate this through community engagement, confidentiality assurances, and validating survey responses with environmental testing.

## Introduction

### Background and Context

Sanitary sewer connections are unavailable for many rural or disadvantaged communities within the United States (1). Residents without access to sewer connections typically rely instead on onsite sewage disposal systems (OSDS) – typically conventional septic systems – though these systems are vulnerable to failure due to inappropriate design, inadequate maintenance, unsuitable soils, high water tables, and climate-related events (2). The *Transforming Wastewater Infrastructure in the United States* project, supported by Columbia World Projects, aims to address these issues by presenting alternative wastewater technology solutions that are suited for use in rural and under-resourced settings. As part of this project, the team is piloting the installation of a community-scale wastewater treatment and collection system in a small rural community located in the Black Belt region of Alabama.

Alabama’s Black Belt is named for the dark, fertile soil that covers 17 counties in central Alabama, including Barbour, Bullock, Butler, Choctaw, Crenshaw, Dallas, Greene, Hale, Lowndes, Macon, Marengo, Montgomery, Perry, Pike, Russell, Sumter, and Wilcox; this is sometimes expanded to include Lamar, Pickens, Bibb, Clarke, Washington, Monroe, Conecuh, and Escambia. While the region is characterized by a rich cultural heritage and is the birthplace of the Civil Rights Movement in the United States, economic development remains limited (3). The state of Alabama consistently ranks among the states with the highest poverty rates, and as of 2023, Alabama had the 8^th^ highest poverty rate in the country (4). The statewide poverty rate is 15.6%, while the average poverty rate of the counties that make up the Black Belt is 24%. Many Black Belt communities experience high poverty, poor health outcomes, and inadequate access to health care (4–6).

Alabama’s Black Belt has recently gained national attention for the region’s failing or nonexistent wastewater infrastructure (7,8). It has been estimated that only 50% of households in the Black Belt are connected to municipal sewer systems, as compared to the national average for the U.S., 75% (9). While municipal sewerage is the ideal strategy for wastewater management in most cases, costs are high, and this model is unworkable where low housing density does not allow for capturing economies of scale. Also, low-income communities already served by conventional municipal sewerage may lack the resources for reliable wastewater treatment and disposal. For example, a recent survey found that more than 1/3 of utility providers across the state of Alabama do not have the means to properly cover the cost of operation, which makes funding updates and maintenance to the systems impossible (10).

Onsite or decentralized sanitation systems are common where conventional sewerage does not reach. Over 40% of Alabamians use an onsite wastewater treatment system, including the majority of those living in the largely rural Black Belt (12). The design of decentralized wastewater systems varies widely, but by far the most common and usually cost-effective model is a conventional septic system. These systems consist of two main elements: a subsurface septic tank, retaining solids that must eventually be emptied, and a drain field that facilitates subsurface discharge of aqueous effluent to the soil. Since wastewater containment depends on keeping effluent discharges below the surface and therefore away from people who could be exposed, systems are designed based on soil permeability (13). When the production of effluent exceeds the capacity of the soil for infiltration, the system fails and wastewater can appear on the surface of the ground, presenting exposure risks to people and the environment. Conventional septic systems perform poorly in much of the Black Belt due to the region’s heavy clay soils that are impermeable to water, though they have been widely implemented. Recent estimates suggest that 40% - 90% of onsite sewage disposal systems in the Black Belt are functioning poorly or completely failing (14). Failing onsite systems can result in sewage back-ups into homes and yards. For instance, after an extreme weather event in 2017, 42% of survey participants in Lowndes County reported having raw sewage backing up into their homes from bathtubs, sinks, and toilets (15). A failing septic system can be expensive to repair or replace; as of 2025, the installation of an alternative system that can safely operate with low-permeability soils can cost up to $30,000 in this region (16). This is not financially feasible for many residents in the Black Belt, where many live at or below the poverty line (17–19).

Without reliable and affordable alternatives, many households use what is known locally as “straight piping.” Straight-piping consists of a pipe that transports untreated wastewater (black water and/or gray water) directly from the home to an uncontained pit, ditch, stream, waste field, or another “disposal area” where it is accessible to people and animals (16). The Alabama Center for Rural Enterprise claims that most straight pipes are less than 10 meters long, suggesting that discharge points are typically within close proximity to homes (15). It is difficult to find data that encapsulates the full scope of this issue since homeowners can face legal repercussions for discharging wastewater via straight pipes. The use of “inadequate sewage disposal systems” is a criminal misdemeanor in Alabama that could result in fines – as much as $500 for each citation – eviction, and – in some cases – arrest (7). Considering the barriers to municipal sewerage and failures of onsite sewage disposal systems and the gaps these create, there is a need for innovation in this space to identify technologies and infrastructure models suitable for rural regions such as the Black Belt.

### Description of Intervention

This protocol details an evaluation of an infrastructure intervention that attempts to overcome the barriers to conventional sewerage and on-site sewage disposal systems by implementing a cluster system approach in a small town in the Alabama Black Belt. While most households are connected to the county water supply, there is no municipal sewerage serving this community. Households will be served by both septic tanks that remove solids from the wastewater and a pump to convey the liquid effluent to a localized treatment system. These systems are typically referred to as a septic tank effluent pump (STEP) or liquid-only sewer. To replace failing septic systems, straight pipes, and other forms of surface discharge of wastewater, households (∼140) within and adjacent to a planned service area will be offered connections to the STEP system under the local Governmental Utility Service Corporation (GUSC).

The infrastructure development is funded through the American Rescue Plan Act (ARPA) with funds allocated from the Alabama Department of Environmental Management to the University of South Alabama. Through this infrastructure project, participating households within this town will receive and be connected to individual onsite tanks (∼1000 gallon) for collection of primary solids, with liquid-only effluent pumped via a small-bore sewer to a localized treatment system (Orenco AX-Max). The Orenco AX-Max is an above ground, modular unit that treats liquid effluent via a recirculating media filter. Treated wastewater effluent from the AX-Max units will be disinfected by ultraviolet radiation (UV) and discharged to a local creek that runs through town. Discharge permits will be obtained from the Alabama Department of Environmental Management (ADEM).

This infrastructure will be implemented across three phases. In Phase I, completed June 2024, one Orenco AX-Max Unit was installed at a university affiliated architecture program with student housing. During Phase II, the system will be expanded to include centralized community spaces and connections will be offered to residences within the town limits. In Phase III, connections will be offered to neighboring, unincorporated communities.

### Intervention Fidelity and Adherence

This infrastructure project is piloting a new approach to wastewater treatment in the Alabama Black Belt and intends to demonstrate that safe, reliable, and effectively managed wastewater services can result in positive environmental and health outcomes for small communities at an affordable cost (capital and operation and maintenance). The evaluation team is conducting an independent assessment of this project and its impacts on the community, and do not benefit from or have influence over the success or failure of the intervention. To assess the fidelity of this intervention, we will collect data on the installation and functionality of both the household septic tank effluent pump systems and the centralized treatment system. We will gather data on household connections and use thorough observational and self-reported sanitary surveys. We will assess the performance of the system through longitudinal environmental sampling at household and community levels.

### Research Objectives and Hypotheses

The evaluation team will examine the effects of the infrastructure project on environmental contamination, infection risk, and individual well-being. These elements are addressed in the following primary (Objective 1) and secondary (Objectives 2-4) research objectives:

<u>Objective 1:</u> Measure the density of fecal microbes (enteric pathogens and fecal indicators) released to the local environment at the household and community levels before and after the implementation of a community-wide sanitation system. <u>Hypothesis:</u> Community-wide sanitation will result in a reduction in the density of enteric pathogens and fecal indicators in the environment.

<u>Objective 2</u>: Collect qualitative and quantitative data regarding the type and functionality of pre- and post-intervention sanitation infrastructure at the household level through participant-taken and observational surveys. <u>Hypothesis:</u> The community-wide sanitation system will result in a reduction in the prevalence of failing sanitation systems and straight-pipe discharge sites.

<u>Objective 3:</u> Model the effects of the implementation of a community-wide sanitation system on the enteric infection risks associated with exposure to enteric pathogens and fecal indicators. <u>Hypothesis:</u> Community-wide sanitation will reduce infection risks associated with exposure to enteric pathogens and fecal indicators.

<u>Objective 4:</u> Explore whether and how the changes in sanitation service affect the quality of life of the participants included in the study. <u>Hypothesis:</u> The intervention will improve the quality of life for participants.

## Methods and analysis

We will collect repeated environmental and household samples (wastewater, soil, drinking water, and surface water) across six structured time points to evaluate changes before and after intervention implementation, as outlined in Figure 1. All samples will be cultured for *E. coli*, and all soil, surface water, and wastewater samples will be analyzed for pre-specified enteric pathogens and fecal markers using quantitative PCR (qPCR). In addition to environmental sampling, structured surveys are conducted at selected time points: a demographic survey at time point one, sanitary inspection surveys at time points two and five, and a well-being survey at time points three and six (Figure 1). Further detail on survey content and analysis is provided in the detailed objectives below. The study began in August 2024 and will conclude at least three months after the final household in Phase II is connected to the intervention, currently targeted for December 2026. Each household follows the same six-point sampling schedule, although the exact dates vary depending on when the intervention reaches each home. *Interrupted Time Series*

To evaluate the effectiveness of this sanitation intervention, we will assess changes in environmental fecal contamination, as indicated by levels of culturable *E. coli* in soil, as our primary trial outcome. Environmental sampling offers a direct measure of how human waste is being contained or released into the environment and is fit to capture the potential health and environmental benefits of improved sanitation infrastructure. While the study includes various ways to evaluate impact (surveys, infrastructure inspections, and qualitative data), this approach allows us to quantify environmental contamination in a way that reflects both function and exposure risk. We focus on *E. coli* as a fecal indicator because it is routinely used to assess sanitary conditions and potential health hazards and based on preliminary data showing elevated culturable *E. coli* counts in impacted soils and surface waters (20–22). We focus on soil as a primary medium of concern given the prevalence of straight pipes and other systems that discharge directly to the surface in this region (9). For the purposes of this study soil also offers a more localized matrix than surface water. Soil will be collected directly from a household’s property and better reflects onsite contamination. Surface water can be intermittent, weather-dependent, and influenced by upstream contamination, making it harder to attribute *E. coli* to a specific source or sanitation system.

We will use an interrupted time series (ITS) design, which allows us to assess both pre-intervention trends and any shifts in *E. coli* concentrations following the introduction of the new improved sanitation infrastructure. In an ITS, longitudinal observations are “interrupted” by an intervention, and observations span the post-intervention period. In the context of sanitation infrastructure expansion, the point of intervention is often staggered as improvements come online and households connect, rather than at a single time point, so participating households “crossover” to the intervention once connected (23). An ITS also includes a counterfactual, the continued projected trend in a hypothetical scenario in which the intervention was not put in place. This provides a comparison for the impact of the intervention by discerning any changes between the observed data and the counterfactual (23,24).

We chose to use an ITS instead of a randomized controlled trial because in the context of community-wide infrastructure projects, randomized trials are infeasible and may come with additional equity and ethical concerns, regardless of the fact that health effects specifically attributable to sanitation are variable across trials (25,26). To address concerns that may arise in the absence of a randomized controlled trial, we will include a minimum of 10 additional households from a neighboring community that has existing adequate sanitation and 10 more comparison households with poor sanitation that will not be connected to the intervention until after the conclusion of this study. These 20 comparison sites will allow estimates of (1) background levels of fecal microbes in sample matrices and (2) the influence of non-intervention-related factors (e.g., flooding) on the presence and quantity of sanitation-related microbes. The comparison sites, along with the fact that we are sampling at multiple time points before and after the intervention, will allow us to account for temporal and spatial variation in microbial density.

### Sample Size Calculations

We calculated a sample size for our primary outcome using a time series regression appropriate for an ITS design. Here we calculate the number of households, *n*, that will be followed longitudinally across the six sampling time points, *t*, (three before and three after the intervention) to detect a meaningful change in soil *E. coli* concentrations with 80% power at a 5% significance level. Equation 1 calculates the crude number of households, *m,* needed before accounting for clustering (27).

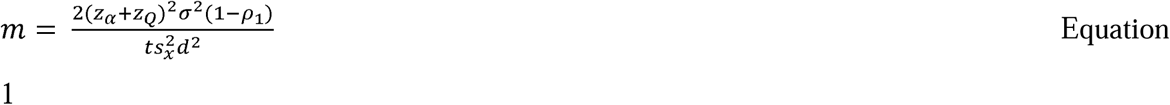

In this equation the term σ*²* captures the total variability in *E. coli* concentrations across all locations and time points, while *s ²* accounts for the variability in repeated measurements within the same location. The intraclass correlation coefficient, ρ_l_reflects how similar those repeated measures are over time for a given household. Finally, the effect size, *d*, is the expected change in log-transformed *E. coli* concentrations in soil attributable to the sanitation intervention.

To estimate the values used in this calculation, we used log-transformed concentrations of *E. coli* ybbW in soil samples based on preliminary data from soil samples collected in Hale County in 2023 (22). For sample size calculations, we assumed that *E. coli* ybbW in soil could serve as a proxy for culturable *E. coli* in soil, based on a moderate positive correlation observed between the two measures in soil samples (Spearman’s ρ = 0.54, *p* = 0.03). The total variance, σ*²*, was calculated as 5.2 log (copies/gram)² based on all available soil samples. The within-household variance, *s ²*, was estimated as 4.3 log (copies/gram)² from a mixed-effects model including a random intercept for location. The intraclass correlation coefficient, p_l_, which reflects the correlation of repeated measurements within a single location over time, was estimated to be 0.16. The effect size, *d*, was defined as a 0.45-log (copies/gram) reduction in soil concentrations, which corresponds to 20% of the standard deviation of log-transformed values from the pilot data. We selected this threshold to reflect a small but meaningful reduction in environmental fecal contamination. Using these inputs and *t* = 6 time points per household, we calculated an intermediate value of *m* = 13 households needed before adjusting for clustering.

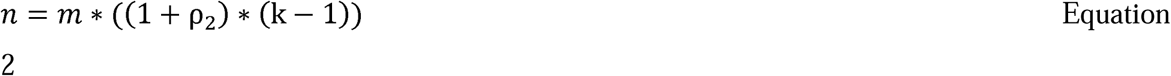

To account for clustering of soil samples within sites, we applied Equation 2 (27). The intracluster correlation coefficient, ρ_2_, used in this adjustment was estimated as 0.17 from a model based on the preliminary data set that included random intercepts for both location and sampling date. We define *k* as the number of observations within each visit—in this case, the total number of soil samples collected for each soil type (impacted versus unimpacted) —which was set to *k* = 3. This adjustment yielded a final, design-corrected sample size of approximately *n* = 30 households needed to detect the specified effect size with 80% power at the 5% significance level.

### Eligibility and Enrollment

We will recruit households through public presentations at publicized community meetings, through the distribution of postcards via mail, door-to-door contact, and at a booth at the local farmer’s market. Recruitment material will include a description of this project, eligibility requirements, and how participants will be compensated for their time and knowledge. Eligible households will be located in the town or in neighboring, unincorporated communities. All households within the town limits will be eligible to be connected to the intervention. At least one person who is 19 years or older - the age of majority in Alabama - and resides at the enrolled household will act as a participant in the study and answer the survey questions. If the primary participant is unavailable at a given time point, another eligible household member may represent the household for that visit, as long as they meet the study requirements and undergo the informed consent process.

### Participant and Public Involvement Statement

Community members will be involved from the beginning of this study, including during the development of research questions, outcome measures, and study design. As previously described, this study will be co-developed with input from local partners including the Auburn Rural Studio and the Black Belt Community Foundation and is guided by a community advisory committee composed of local residents and community leaders. The local partners and community advisory committee provided feedback on the survey content and overall study approach to ensure alignment with community priorities. The advisory committee reviewed study materials to assess the burden of participation and helped make sure the study design was respectful of all participants’ time and privacy. Community partners have also supported recruitment efforts by mailing flyers and organizing local outreach events. Community partners and leaders will be actively involved in the dissemination of results, including input on decisions on what findings to share, in what format, and when.

## Objective 1

Measure the density of fecal microbes released to the local environment (enteric pathogens and fecal indicators) at the household and community levels before and after the implementation of a community-wide sanitation system.

### Overview of the Study Design

We are collecting longitudinal microbial density data from environmental and household samples to estimate the reduction of pathogens and fecal markers released into the environment following the expansion of the wastewater treatment intervention. From the 50 recruited households, we will collect soil, wastewater, and drinking water samples to analyze for fecal indicators and enteric pathogens. We will conduct longitudinal surface water sampling from the receiving waters upstream and downstream of the discharge point of the new wastewater treatment system. All community members—both participants and non-participants—will be made aware that community-level sampling will be taking place for the purposes of this study at the community meetings described previously.

We will culture fecal indicator bacteria (total coliforms and *E. coli*) from environmental and household samples (wastewater, soil, drinking water, and surface water) using the IDEXX Colilert-18 Quanti-Tray 2000 System as independent predictors of exposure risk, to develop pathogen:indicator ratios for use in risk modeling, and as proxy indicators of pathogen viability in samples. We will further analyze the soil, wastewater, and surface water samples for pre-specified pathogens and fecal markers by real-time quantitative PCR (qPCR) using a custom TaqMan Array Card (28). Targets are specified in Appendix 1. We will use the microbial data collected before and after connection to the intervention to inform an interrupted time series (ITS) analysis. We are particularly interested in assessing the impact of the intervention on pathogens in the environment through drinking water, surface water, and soil sampling as proximal, objective, quantitative measures on the causal pathway from the intervention to all downstream health and non-health effects of improved sanitation services.

### Drinking Water Sample Collection

We will collect drinking water samples from all 50 households at each of the six time points. All of the households will have a 1000 mL and 50 mL grab sample of their drinking water collected. This grab sample will be used for analysis as outlined above.

### Soil Sample Collection

At each of the six time points, three soil samples will be collected from an area thought to be impacted by failing sanitation, as identified by the participant living in the household. If the household is relying on a cesspool or straight pipe, a sample will be collected directly at the point of impact by collecting the top layer of the receiving soil, with additional samples collected one foot away and three feet away from the point of impact. If the household is relying on a septic tank with a septic field, soil samples will be collected from three randomly selected locations overlaying the septic field. Impacted soil samples will not be collected at the 10 households with existing adequate sanitation services, which, by definition, have no directly impacted soils. As a control, three additional soil samples will be collected at all 50 households during each of the six sampling time points in a section of the yard that is identified by the participant as unimpacted by failing sanitation. If no such area exists, then the researcher will randomly sample at three locations at the front, side, and back of the home. Once initially selected, the same household soil locations will be sampled at each of the six time points.

### Wastewater Sample Collection

At each of the six time points, one wastewater sample will be collected at the household’s wastewater receiving site. The sample collected will depend on the participant’s current sanitation infrastructure status. For example, wastewater samples from households that utilize a septic tank will be collected directly from the tank. For households that utilize straight pipes or cesspools, a mixture of surface water, gray water, and human waste will be collected from the discharge site. Sampling directly from the pipe itself will not be possible because flow only occurs during use, such as flushing the toilet, which would be difficult to time and any sample obtained may reflect waste from one individual at one time point. Sampling from the receiving environment will provide a more representative sample of waste discharges and environmental impact. Households that are connected to the municipal wastewater treatment system will not have wastewater samples collected at the household; instead, researchers will collect wastewater influent from the municipal system three randomly selected times during the bounds of each time point. These samples will allow for estimation of pathogen loading to the environment under various containment or discharge scenarios.

### Sample Collection from Receiving Stream

Surface water samples will be collected from two creeks at publicly accessible points upstream and downstream of the town serviced by the intervention and at the future effluent discharge point. These samples will be collected during each environmental sampling time point, which refers to a defined sampling period during which all enrolled households are sampled. After each household is connected to the intervention, 15 samples per time point will be taken of the treatment system influent, 15 samples per time point will be taken at the treatment system effluent prior to UV disinfection, and 15 samples per time point will be taken at the treatment system effluent after UV disinfection.

### Data Analysis and Outcomes

Our primary outcome is to compare the concentration of culturable *E. coli* in the environment pre- and post-intervention using a time series regression analysis suitable for an interrupted time series ITS design. We will use the Durbin-Watson statistic to test the presence of autocorrelation in *E. coli* concentrations over time and will adjust for autocorrelation if found (29). To determine regression estimates that express the level (the average value of *E. coli* concentration at a given time point) and trend (the direction and rate of change in *E. coli* concentrations over time) of the outcomes pre- and post-intervention, we will use the generalized least-squares method with the Prais-Winston estimator (29). Regression coefficients corresponding to standard effect sizes on culturable *E. coli* soil concentrations will be obtained for a step change and a change in trend pre- and post-intervention. A step change, also known as a change in level, is the difference between the observed level at the first intervention time point and the predicted time trend. A change in trend is defined as the difference between the post-intervention and pre-intervention slopes, where slopes are defined as the rate of change in *E. coli* concentration over time (30).

Our secondary outcomes are the estimated prevalence of individual pre-specified pathogens in soil, wastewater, and surface water samples. We will graph the mean and standard deviation scores (the average measured values) for fecal markers and composite measurement, which summarizes the presence or concentration of multiple enteric pathogens within a sample. Within each sample type, only markers and composite indicators appearing in at least 10% of the samples collected at each time point will be included, in order to visualize the trends in fecal indicator concentrations and pathogen prevalence.

## Objective 2

Collect qualitative and quantitative data regarding the type and functionality of existing sanitation infrastructure at the household level through a participant-administered and observational survey.

### Demographic Survey

The team will conduct a demographic survey in person during the first household visit for sampling. The demographic survey will be self-administered by a participant at each of the 50 households unless the participant specifies otherwise. This survey is intended to gather information about the characteristics of the enrolled participant and the household as well as contact information for future sampling and surveys. Each survey will be administered through Qualtrics (https://unc.az1.qualtrics.com/) and will include multiple-choice and open-ended questions. Questions will include but not be limited to the participant’s gender and race/ethnicity, household income range, and preferred method of future contact.

### Sanitary Inspection Survey

The team will conduct observational and participant-taken sanitary inspection surveys in person or over the phone pre- and post-intervention. The observational survey will be conducted at all 50 households by the enumerator and is intended to elucidate current management practices for wastewater at the home as well as to document any visual damages to existing infrastructure. The written participant sanitation surveys will be self-administered by a participant at each of the 50 households unless the participant notes otherwise and are intended to gather information about each household’s water and sanitation practices. Each survey will be administered through Qualtrics (https://unc.az1.qualtrics.com/) and will include multiple-choice questions, open-ended questions, and photographs of current infrastructure. Questions will include where gray and black water is discharged, damage to infrastructure, and what type of water is used for drinking and hygiene.

### Coding and Analysis

We will code responses to the survey questions into appropriate categories and then import data into R for analysis (R Foundation for Statistical Computing, Vienna, Austria). The category codes that will be applied to the responses will be created deductively. Data collected in the surveys will be used to inform the characteristics of the participants of this study. The outcome for objective 2 is a characterization of the wastewater intervention’s potential impacts, such as how human waste is being treated and the observable quality of the household water supply.

## Objective 3

Estimate the effects of the implementation of a community-wide sanitation system on the health risks associated with exposure to enteric pathogens and fecal indicators.

### Quantitative Microbial Risk Assessment Model

We will use a quantitative microbial risk assessment (QMRA) model to compare the annual risk of enteric infection between pre- and post-intervention settings. QMRA is a probabilistic modeling approach that combines pathogen-specific dose-response models and population-specific exposure assessments to characterize the risk of infection from microbial hazards (31–34).

We will estimate the risk of enteric infection under pre- and post-intervention settings using the data collected in Objective 1. From this dataset, we will use measures of prevalence of enteric pathogens and concentrations of pathogen gene targets to estimate a distribution of pathogens in soil under both pre- and post-intervention settings.

We will estimate the viability of the enteric pathogens using a ratio of culturable *E. coli* to gene copies of *E. coli* recovered from the environmental samples. Using the ingestion parameters for soil from the US EPA exposure factors handbook, we will estimate a daily exposure dose of viable enteric pathogens. Using published dose-response models, we will then estimate a daily and annual risk of infection for pathogens that are in more than 10% of the samples. We will use stochastic methods to propagate variability and uncertainty throughout the model (35). We will then use previously described methods to generate effect measures estimating attributable reductions in risk of infection between pre- and post-intervention settings (36). The outcome for objective 3 is a modeled estimate of the change in daily and annual risk of enteric infection due to environmental exposure through soil by comparing pre- and post-intervention soil conditions.

## Objective 4

Explore whether and how the changes in sanitation service affect the quality of life of the participants included in the study.

### Well-Being Survey

We will collect data on participant well-being through a survey. The team will conduct surveys with a participant representing each of the 50 households enrolled in this study. The surveys will be conducted either in person or by phone at two time points, once before and once after the implementation of the community-wide sanitation system. These surveys will attempt to capture how limited access to sanitation services affects the overall well-being such as distress, fear of legal ramifications, and management related concerns. The surveys will be administered through Qualtrics (https://unc.az1.qualtrics.com/) through a combination of multiple-choice and open-ended questions. If the participant consents, the well-being survey will be recorded and transcribed. The outcome for Objective 4 is an assessment of changes in participants’ quality of life by comparing well-being before and after connection to the wastewater intervention.

## Limitations

### Secular trends in exposures or outcomes

Because ITS analyses compare pre- and post-intervention trends of outcome measures, a limitation of this approach is the existence of non-intervention, secular changes that obscure or complicate the attribution of effects to the point of intervention. Additionally, an ITS does not include a control group; therefore, simultaneous trends may be incorrectly attributed to the intervention itself. Weather-related events, such as rainfall and temperature, are examples of independent variables that could act as potential confounders because extreme rainfall or temperatures can increase blockages and breakages in onsite wastewater treatment systems (37). We will closely monitor rainfall and adjust our analyses to account for any differences observed by incorporating comparison sites. However, it is important to note that the potential for residual confounding cannot be eliminated completely in this study design. As previously described, we plan to include a minimum of 10 comparison sites with existing adequate sanitation and 10 comparison sites with poor sanitation that will not be connected to the intervention until after this study concludes. This plan will allow estimates of (1) background levels of fecal microbes in sample matrices and (2) the influence of non-intervention-related factors (e.g., flooding) on the presence and quantity of sanitation-related microbes.

### Bias

There are several possible sources of bias in this study design. We are specifying microbial targets *a priori*, limiting analysis of samples to a pre-selected list of fecal indicators and enteric pathogens. We are attempting to minimize this bias by basing our TAC design on baseline data from a series of studies before the impact assessment. Another form of possible bias is reporting bias in the surveys and interviews. To attempt to minimize reporting bias, we are pairing this data with the independent quantitative environmental sample data collected.

Additionally, there is potential for volunteer or self-selection bias, as participation in this study is not required for households connecting to the system. This may bias data by only including people who would typically volunteer to be a part of a study and/or those who have enough time to participate in research activities. Similarly, participants must be willing to pay a monthly maintenance fee to connect to the intervention (at the time of this writing, the exact pricing for tariffs or connection fees have not been established), which could select for more affluent participants while excluding those unable to afford the fee. It is also possible that households with the most inadequate or failing sanitation systems may be more motivated to participate, seeing greater potential benefit from the intervention. Furthermore, there is a chance that we will experience courtesy bias, which occurs when participants give overly positive responses in surveys or interviews to avoid offending the researcher or to present themselves favorably. We will attempt to minimize the amount of courtesy bias in this study by working with the community advisory group, being very clear on how this data will be used, and making sure it is known that this team is working for the community and is not affiliated with nor invested in the intervention itself. Finally, we predict that we will also experience social desirability bias, which is when participants respond to certain questions in a way that they believe will positively portray them in line with social norms. We will attempt to minimize the amount of social desirability bias in this study by ensuring participants that their responses will be kept completely confidential and anonymous.

## Plan for Delayed or Failed Implementation of the Intervention

While this study is based on the implementation of the wastewater intervention, we recognize the possibility that delays or failures in delivery could occur. If the intervention is not completed according to the planned timeline, and no households are connected by December 2026, we will take several steps to ensure the scientific and community value of this research effort is maintained as much as possible. All pre-intervention sampling, surveys, and analysis will be completed regardless of the status of implementation, and these data will still support important scientific insights. First, we will conduct a model-based microbial risk assessment using the collected environmental data from pre-intervention sampling to estimate potential exposure reductions had the intervention occurred as planned. Second, we will conduct a structured analysis to understand why the intervention did not move forward, including institutional, logistical, and financial barriers, incorporating both quantitative and qualitative methods to assess community readiness, planning barriers, and environmental baselines. Third, we will conduct in-depth interviews with key stakeholder groups to better understand the infrastructural, institutional, and social constraints that may have delayed or prevented implementation.

## Ethics and dissemination

Informed consent will be obtained for all participants prior to any data collection. During informed consent, participants will be provided with a clear explanation of the purpose of the study, sampling and survey procedures, risks, benefits, confidentiality protections, and how study results will be shared. Given the sensitive nature of household sanitation practices, participants in this study will be informed and reminded throughout the study of their rights and that participation is completely voluntary. Additionally, all identifiable data will be stored separately from all other data and protected, using secure password-protected servers. Deidentified study results will be shared with all participants and residents of the community during a community forum hosted in a public location. Upon request, each household will receive a summary of their site-specific data. Deidentified study results will be shared in peer-reviewed publications. Supporting documentation and deidentified datasets will be made available for public use in line with the best practices of open science.

## Data Availability

Upon publication of results for this study, the household data will be fully deidentified and made freely available in a permanent online repository (https://osf.io/9kvfa/).

## Acknowledgements

We are deeply grateful to the community members who generously welcomed us and made this project possible through their time and trust.

## Author Contributions

Olivia A. Harmon designed the protocol and wrote the initial draft of this manuscript. Megan E.J. Lott and David Holcomb contributed to the study design, especially as it related to environmental microbiological sampling and analysis. Mark Elliott and Kevin White conceived the plan for the implementation of the intervention and greatly assisted with the description of the intervention. Emily McGlohn and Christopher H. Spencer contributed to the study design, especially as it related to community outreach. As the principal investigator, Joe Brown oversaw the design of the study and the writing of this protocol. Olivia A. Harmon, Megan E.J. Lott, and Joe Brown are a part of the evaluation team. All authors contributed to the final draft of this manuscript and approved it for publication.

---

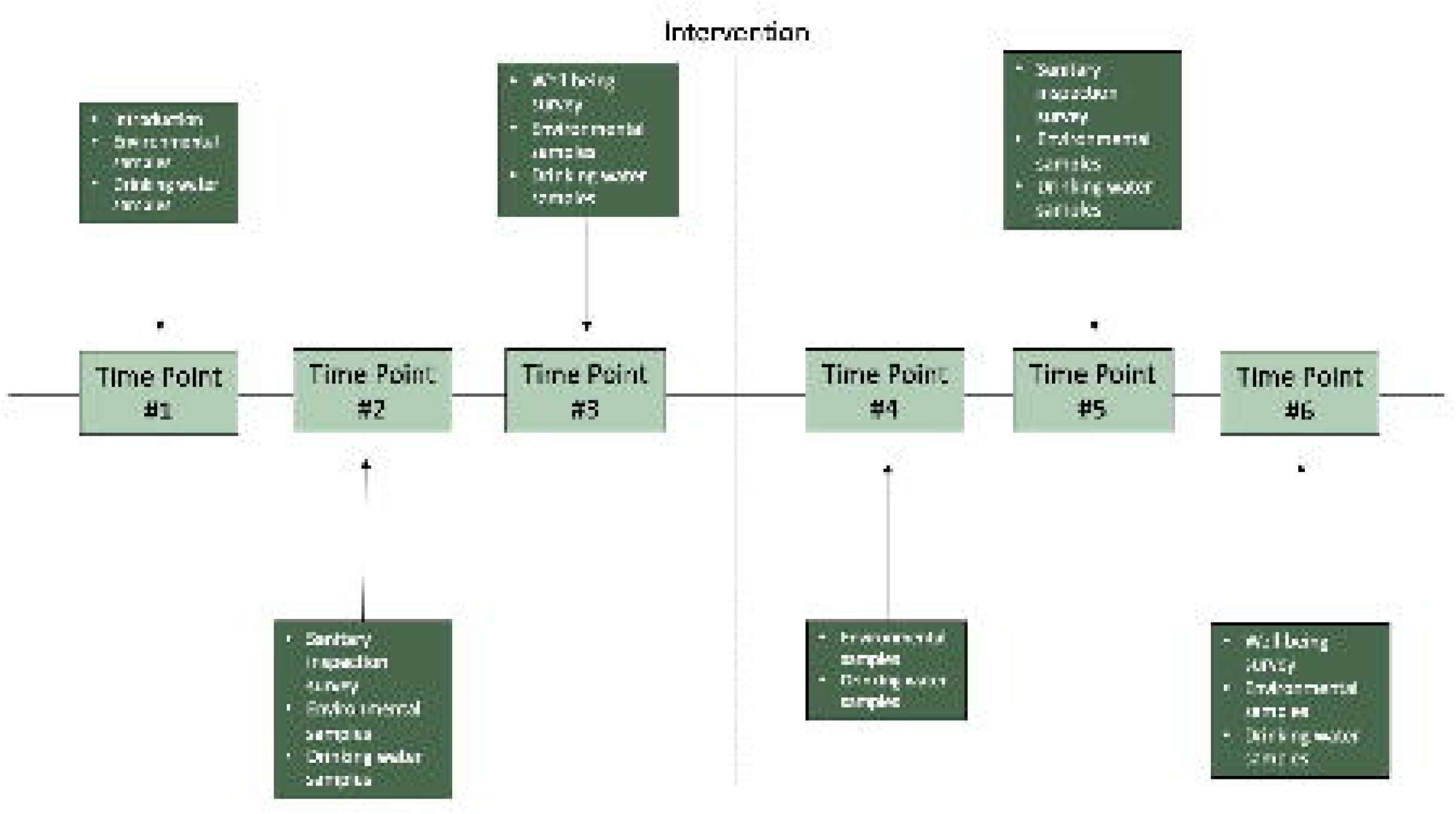

## References

1. Flowers CC, Ward JK, Winkler IT. Flushed and Forgotten: Sanitation and Wastewater in Rural Communities in the US | Institute for the Study of Human Rights. [cited 2022 Nov 17]; Available from: https://www.humanrightscolumbia.org/news/flushed-and-forgotten-sanitation-and-wastewater-rural-communities-us

2. Maxcy-Brown J, Elliott MA, Krometis LA, Brown J, White KD, Lall U. Making waves: Right in our backyard-surface discharge of untreated wastewater from homes in the United States. Water Res. 2021 Feb;190:116647.

3. Jeffries HK. Bloody Lowndes: Civil Rights and Black Power in Alabama’s Black Belt [Internet]. Bloody Lowndes. New York University Press; 2009 [cited 2022 Nov 16]. Available from: https://www.degruyter.com/document/doi/10.18574/nyu/9780814743652.001.0001/html

4. Poverty in the United States: Explore the Map [Internet]. Center for American Progress. 2024 [cited 2025 Jun 23]. Available from: https://www.americanprogress.org/data-view/poverty-data/poverty-data-map-tool/

5. Katsinas SG, Till GA, Peterson J, Keeney NE, Kelly PJ, Bray NJ. Halfway Home and a Long Way To Go: Bridging Persistent Poverty Gap in Alabama’s Black Belt.

6. Jacobs E, Whann H, Corley EG, Bowen J, Keeney N. Healthcare: A Key Challenge in Alabama’s Black Belt.

7. Okeowo A. The Heavy Toll of the Black Belt’s Wastewater Crisis. The New Yorker [Internet]. 2020 Nov 23 [cited 2022 Nov 20]; Available from: https://www.newyorker.com/magazine/2020/11/30/the-heavy-toll-of-the-black-belts-wastewater-crisis

8. Office of Public Affairs | Departments of Justice and Health and Human Services Announce Interim Resolution Agreement in Environmental Justice Investigation of Alabama Department of Public Health | United States Department of Justice [Internet]. 2023 [cited 2023 Dec 12]. Available from: https://www.justice.gov/opa/pr/departments-justice-and-health-and-human-services-announce-interim-resolution-agreement

9. Maxcy-Brown J, Wilson T, Chai R, McCaskill H, Bakchan A, Christian L, et al. The Past, Present, and Future of Wastewater Management in Alabama’s Black Belt. J Sustain Water Built Environ. 2024 Nov 1;10(4):04024007.

10. Hall T. Assisting Our Black Belt Neighbors: Third World Problems in Our Backyard.

11. He J, Dougherty M, Zellmer R, Martin G. Assessing the Status of Onsite Wastewater Treatment Systems in the Alabama Black Belt Soil Area. Environ Eng Sci. 2011 Oct;28(10):693–9.

12. US EPA O. Onsite Wastewater Treatment and Disposal Systems [Internet]. 2015 [cited 2022 Nov 21]. Available from: https://www.epa.gov/septic/onsite-wastewater-treatment-and-disposal-systems

13. US EPA O. Types of Septic Systems [Internet]. 2018 [cited 2024 May 27]. Available from: https://www.epa.gov/septic/types-septic-systems

14. Williams MK, Murthy S. Report on the October 25, 2011 Roundtable Meeting with the UN Special Rapporteur on the Human Right to Safe Drinking Water and Sanitation, Catarina De Albuquerque. In: 2011 Roundtable Meeting with the UN Special Rapporteur on the Human Right to Safe Drinking Water and Sanitation, Catarina De Albuquerque (October 2011). 2011.

15. McKenna ML, McAtee S, Bryan PE, Jeun R, Ward T, Kraus J, et al. Human Intestinal Parasite Burden and Poor Sanitation in Rural Alabama. Am J Trop Med Hyg. 2017 Nov 8;97(5):1623–8.

16. Izenberg M, Johns-Yost O, Johnson PD, Brown J. Nocturnal Convenience: The Problem of Securing Universal Sanitation Access in Alabama’s Black Belt. Environ Justice. 2013 Dec;6(6):200–5.

17. Katsinas SG, Till G, Corley EG, O’Brien S, Courchesne E, Bray N. Poverty, Housing, & GDP in Alabama’s Black Belt. :22.

18. Bureau UC. Census.gov. [cited 2022 Nov 20]. Poverty in the United States: 2021. Available from: https://www.census.gov/library/publications/2022/demo/p60-277.html

19. alabama poverty - Census Bureau Tables [Internet]. [cited 2022 Nov 20]. Available from: https://data.census.gov/table?q=alabama+poverty&g=0100000US

20. Navab-Daneshmand T, Friedrich MND, Gächter M, Montealegre MC, Mlambo LS, Nhiwatiwa T, et al. Escherichia coli Contamination across Multiple Environmental Compartments (Soil, Hands, Drinking Water, and Handwashing Water) in Urban Harare: Correlations and Risk Factors. Am J Trop Med Hyg. 2018 Mar;98(3):803–13.

21. Pickering AJ, Ercumen A, Arnold BF, Kwong LH, Parvez SM, Alam M, et al. Fecal Indicator Bacteria along Multiple Environmental Transmission Pathways (Water, Hands, Food, Soil, Flies) and Subsequent Child Diarrhea in Rural Bangladesh. Environ Sci Technol. 2018 Jul 17;52(14):7928–36.

22. Harmon OA, Lott MEJ, Elliott M, McGlohn E, Brown J. Environmental pathogen hazards reveal need for improved sanitation infrastructure in Alabama’s Black Belt. 2025 May 29 [cited 2025 Jun 23]; Available from: https://eartharxiv.org/repository/view/9292/

23. Lopez Bernal J, Cummins S, Gasparrini A. Interrupted time series regression for the evaluation of public health interventions: a tutorial. Int J Epidemiol. 2016 Jun 9;dyw098.

24. Shadish, WIlliam R., Cook, Thomas D., Campbell, Donald T. Experimental and Quasi-Experimental Designs for Generalized Causal Inference.

25. Trials and Tribulations: The Rise and Fall of the RCT in the WASH Sector | Randomized Control Trials in the Field of Development: A Critical Perspective | Oxford Academic [Internet]. [cited 2023 Dec 5]. Available from: https://academic.oup.com/book/31966/chapter/267702976

26. Arnold BF, Khush RS, Ramaswamy P, London AG, Rajkumar P, Ramaprabha P, et al. Causal inference methods to study nonrandomized, preexisting development interventions. Proc Natl Acad Sci U S A. 2010 Dec 28;107(52):22605–10.

27. Diggle P. Analysis of Longitudinal Data. OUP Oxford; 2002. 396 p.

28. Lappan R, Henry R, Chown SL, Luby SP, Higginson EE, Bata L, et al. TaqMan Array Cards enable monitoring of diverse enteric pathogens across environmental and host reservoirs [Internet]. bioRxiv; 2020 [cited 2023 Feb 2]. p. 2020.10.27.356642. Available from: https://www.biorxiv.org/content/10.1101/2020.10.27.356642v1

29. Durbin J. Testing for Serial Correlation in Least-Squares Regression When Some of the Regressors are Lagged Dependent Variables. Econometrica. 1970 May;38(3):410.

30. Ramsay CR, Matowe L, Grilli R, Grimshaw JM, Thomas RE. INTERRUPTED TIME SERIES DESIGNS IN HEALTH TECHNOLOGY ASSESSMENT: LESSONS FROM TWO SYSTEMATIC REVIEWS OF BEHAVIOR CHANGE STRATEGIES. Int J Technol Assess Health Care. 2003 Dec;19(4):613–23.

31. Brouwer AF, Masters NB, Eisenberg JNS. Quantitative Microbial Risk Assessment and Infectious Disease Transmission Modeling of Waterborne Enteric Pathogens. Curr Environ Health Rep. 2018 Jun;5(2):293–304.

32. Moazeni M, Nikaeen M, Hadi M, Moghim S, Mouhebat L, Hatamzadeh M, et al. Estimation of health risks caused by exposure to enteroviruses from agricultural application of wastewater effluents. Water Res. 2017 Nov 15;125:104–13.

33. Mok HF, Barker SF, Hamilton AJ. A probabilistic quantitative microbial risk assessment model of norovirus disease burden from wastewater irrigation of vegetables in Shepparton, Australia. Water Res. 2014 May 1;54:347–62.

34. Soller JA, Eftim SE, Nappier SP. Direct potable reuse microbial risk assessment methodology: Sensitivity analysis and application to State log credit allocations. Water Res. 2018 Jan;128:286–92.

35. Haas CN, Rose JB, Gerba CP. Quantitative Microbial Risk Assessment. John Wiley & Sons; 2014. 439 p.

36. Capone D, Bivins A, Brown J. Producing ratio measures of effect with quantitative microbial risk assessment. Risk Anal. 2023 May;43(5):917–27.

37. Hughes J, Cowper-Heays K, Olesson E, Bell R, Stroombergen A. Impacts and implications of climate change on wastewater systems: A New Zealand perspective. Clim Risk Manag. 2021 Jan 1;31:100262.

