## Appendix 1 for "A study protocol for an interrupted time series analysis and pre-post surveys to assess the effects a community-wide sanitation system on environmental contamination, infection risk, and well-being in Alabama’s Black Belt"

| **Pathogen** | **Forward Sequence(s) (5'-3')** | **Reverse Sequence(s) (5'-3')** | **Probe Sequence (5'-3')** | **Source** |
| --- | --- | --- | --- | --- |
| 16S | CCTACGGGDGGCWGCA | GGACTACHVGGGTMTCTAATC | CAGCAGCCGCGGTA | (1) |
| 18S (Manufacturer's Controls) |  |  |  |  |
| *Acanthamoeba* spp. | CCCAGATCGTTTACCGTGAA | TAAATATTAATGCCCCCAACTATCC | CTGCCACCGAATACATTAGCATGG | (2) |
| Adenovirus 40/41 | AACTTTCTCTCTTAATAGACGCC | AGGGGGCTAGAAAACAAAA | CTGACACGGGCACTCT | (3) |
| *Ancylostoma duodenale* | GAATGACAGCAAACTCGTTGTTG | ATACTAGCCACTGCCGAAACGT | ATCGTTTACCGACTTTAG | (3) |
| *Ascaris lumbricoides* | GTAATAGCAGTCGGCGGTTTCTT | GCCCAACATGCCACCTATTC | TTGGCGGACAATTGCATGCGAT | (4) |
| *Astroviridae* | CAGTTGCTTGCTGCGTTCA | CTTGCTAGCCATCACACTTCT | CACAGAAGAGCAACTCCATCGC | (3) |
| *Balantidium coli* | TGCAATGTGAATTGCAGAACC | TGGTTACGCACACTGAAACAA | CTGGTTTAGCCAGTGCCAGTTGC | (5) |
| *Blastocystis* spp. | TGGTCCGRTGAACACTTTGGAT | CCTACGGAAACCTTGTTACGACTTCA | CTTCCTCTAAATGRTAAGATT | (3) |
| Bovine herpesvirus (BHV) | TGTGGACCTAAACCTCACGGT | GTAGTCGAGCAGACCCGTGTC | AGGACCGCGAGTTCTTGCCGC | (6) |
| Bovine respiratory syncytial virus (BRSV) | GCAATGCTGCAGGACTAGGTATAAT | ACACTGTAATTGATGACCCCATTCT | ACCAAGACTTGTATGATGCTGCCAAAGCA | (7) |
| *Campylobacter jejuni*/*coli* | CTGCTAAACCATAGAAATAAAATTTCTCAC | CTTTGAAGGTAATTTAGATATGGATAATCG | CATTTTGACGATTTTTGGCTTGA | (8) |
| *Clostridioides difficile* | GGTATTACCTAATGCTCCAAATAG | TTTGTGCCATCATTTTCTAAGC | CCTGGTGTCCATCCTGTTTC | (9) |
| *Cryptosporidium* spp. | GGGTTGTATTTATTAGATAAAGAACCA | AGGCCAATACCCTACCGTCT | TGACATATCATTCAAGTTTCTGAC | (9) |
| *E. coli* gadA/B | GGATATCGTCTGGGACTTCCG | GCGGAGCCAGACCGAATTT | GTGAAATCGATCAGTGCTTCAGGCCA | (10) |
| *E. coli* O157:H7/H-serotype | CAGTCTGGATCGCGAAAACTG | ACCAGACGTTGCCCACATAATT | ATTGAGCAGCGTTGG | (11) |
| *E. coli* ybbW | TGATTGGCAAAATCTGGCCG | GAAATCGCCCAAATCGCCAT | CCGCCG[ZEN]AAAACGATATAGATGCACGG | (12) |
| *Entamoeba histolytica* | ATTGTCGTGGCATCCTAACTCA | GCGGACGGCTCATTATAACA | TCATTGAATGAATTGGCCATTT | (13) |
| Enteroaggregative *E. coli* aaiC | ATTGTCCTCAGGCATTTCAC | ACGACACCCCTGATAAACAA | TAGTGCATACTCATCATTTAAG | (3) |
| Enteroaggregative *E. coli* aatA | CTGGCGAAAGACTGTATCAT | TTTTGCTTCATAAGCCGATAGA | TGGTTCTCATCTATTACAGACAGC | (3) |
| *Enterobius vermicularis* | CAAACAACTGCATCACCAATAAC | AGTGTAGAGCAATAAGCAGTAAAG | TACCAACAACACTTGCACGTCTCTTCA | (14) |
| Enteropathogenic *E. coli* bfpA+ | TGGTGCTTGCGCTTGCT | CGTTGCGCTCATTACTTCTG | CAGTCTGCGTCTGATTCCAA | (9) |
| Enteropathogenic *E. coli* eae+ | CATTGATCAGGATTTTTCTGGTGATA | CTCATGCGGAAATAGCCGTTA | ATACTGGCGAGACTATTTCAA | (9) |
| Enterotoxigenic *E. coli* heat-labile | TTCCCACCGGATCACCAA | CAACCTTGTGGTGCATGATGA | CTTGGAGAGAAGAACCCT | (15) |
| Enterotoxigenic *E. coli* STh | GCTAAACCAGYAGRGTCTTCAAAA | CCCGGTACARGCAGGATTACAACA | TGGTCCTGAAAGCATGAA | (9) |
| Enterotoxigenic *E. coli* STp | TGAATCACTTGACTCTTCAAAA | GGCAGGATTACAACAAAGTT+E5 | TGAACAACACATTTTACTGCT | (9) |
| *Giardia* spp. | GACGGCTCAGGACAACGGTT | TTGCCAGCGGTGTCCG | CCCGCGGCGGTCCCTGCTAG | (13) |
| *Helicobacter pylori* | GACACCAGAAAAAGCGGCTA | AGCGCATGTCTTCGGTTAAA | TCACTAAAGCGTTTTCTACC | (3) |
| Hepatitis A | TCACCGCCGTTTGCCTAG | GGAGAGCCCTGGAAGAAAG | TTAATTCCTGCAGGTTCAGG | (16) |
| Hepatitis G | CGGCCAAAAGGTGGTGGATG | CGACGAGCCTGACGTCGGG | AGGTCCCTCTGGCGCTTGTGGCGAG | (17) |
| Human Enterovirus | 1: CACYGAACCAGARGAAGCCA  2: CACTGAACCAGAGGAAGCTA | 1: CCAAAGCTGCTCTACTGAGAAA  2: CTAAAGCTGCCCTACTAAGRAA | TCGCACAGTGATAAATCARCAYGG | (18) |
| Human Mitochondrial DNA | CAATGAATCTGAGGAGGCTAC | CGTGCAAGAATAGGAGGTG | ACCCTCACACGATTCTTTACCTTTCACT | (19) |
| Human-specific HF183 Bacteroides 16S rRNA genetic marker | ATCATGAGTTCACATGTCCG | CTTCCTCTCAGAACCCCTATCC | CTAATGGAACGCATCCC | (20) |
| *Necator americanus* | CTGTTTGTCGAACGGTACTTGC | ATAACAGCGTGCACATGTTGC | CTGTACTACGCATTGTATAC | (4) |
| Norovirus GI | CGYTGGATGCGNTTYCATGA | CTTAGACGCCATCATCATTYAC | TGGACAGGAGATCGC | (3) |
| Norovirus GII | CARGARBCNATGTTYAGRTGGATGAG | TCGACGCCATCTTCATTCACA | TGGGAGGGCGATCGCAATCT | (3) |
| *Plesiomonas shigelloides* | CCGCCGTGAAGGCAAAG | GCTACCGGCTCACCCAGAT | CACACCCAAGAATAC | (3) |
| Poliovirus | AGCACTTCTGTTTCCC | ACGGACACCCAAAGTA | ACATAAGAATCCTCCGGCCCCTGA | (21) |
| Rotavirus | ACCATCTWCACRTRACCCTCTATGAG | GGTCACATAACGCCCCTATAGC | AGTTAAAAGCTAACACTGTCAAA | (22) |
| *Salmonella enterica* | CTCACCAGGAGATTACAACATGG | AGCTCAGACCAAAAGTGACCATC | CACCGACGGCGAGACCGACTTT | (23) |
| Sapovirus I, II, IV, V | 1: GAYCASGCTCTCGCYACCTAC  2: TTGGCCCTCGCCACCTAC | CCCTCCATYTCAAACACTA | CCRCCTATRAACCA | (24) |
| SARS-CoV-2 | GACCCCAAAATCAGCGAAAT | TCTGGTTACTGCCAGTTGAATCTG | ACCCCGCATTACGTTTGGTGGACC | (25) |
| Shiga toxin producing *E. coli* Stx1 | ACTTCTCGACTGCAAAGACGTATG | ACAAATTATCCCCTGWGCCACTATC | CTCTGCAATAGGTACTCCA | (9) |
| Shiga toxin producing *E. coli* Stx2 | CCACATCGGTGTCTGTTATTAACC | GGTCAAAACGCGCCTGATAG | TTGCTGTGGATATACGAGG | (9) |
| *Shigella* spp. and enteroinvasive *E. coli* | CCTTTTCCGCGTTCCTTGA | CGGAATCCGGAGGTATTGC | CGCCTTTCCGATACCGTCTCTGCA | (26) |
| *Strongyloides stercoralis* | TCCAGAAAAGTCTTCACTCTCCAG | TGCGTTAGAATTTAGATATTATTGTTGCT | TCAGCTCCAGTTGAACAACAGCCTCCAA | (3) |
| *Trichuris trichiura* | TTGAAACGACTTGCTCATCAACTT | CTGATTCTCCGTTAACCGTTGTC | CGATGGTACGCTACGTGCTTACCATGG | (9) |
| *Yersinia enterocolitica* | TGATTCACCAGCAGCAATAC | GGCATCATGAAAGGCGG | TGTCGGTTTCTCCTTCCAGG | (3) |

1.	Liu CM, Aziz M, Kachur S, Hsueh PR, Huang YT, Keim P, et al. BactQuant: an enhanced broad-coverage bacterial quantitative real-time PCR assay. BMC Microbiol. 2012 Apr 17;12:56.

2.	Qvarnstrom Y, Visvesvara GS, Sriram R, da Silva AJ. Multiplex real-time PCR assay for simultaneous detection of Acanthamoeba spp., Balamuthia mandrillaris, and Naegleria fowleri. J Clin Microbiol. 2006 Oct;44(10):3589–95.

3.	Liu J, Gratz J, Amour C, Nshama R, Walongo T, Maro A, et al. Optimization of Quantitative PCR Methods for Enteropathogen Detection. PLOS ONE. 2016 Jun 23;11(6):e0158199.

4.	Basuni M, Muhi J, Othman N, Verweij JJ, Ahmad M, Miswan N, et al. A Pentaplex Real-Time Polymerase Chain Reaction Assay for Detection of Four Species of Soil-Transmitted Helminths. Am J Trop Med Hyg. 2011 Feb 4;84(2):338–43.

5.	Sow D, Parola P, Sylla K, Ndiaye M, Delaunay P, Halfon P, et al. Performance of Real-Time Polymerase Chain Reaction Assays for the Detection of 20 Gastrointestinal Parasites in Clinical Samples from Senegal. Am J Trop Med Hyg. 2017 Jul 12;97(1):173–82.

6.	Wang J, O’keefe J, Orr D, Loth L, Banks M, Wakeley P, et al. Validation of a real-time PCR assay for the detection of bovine herpesvirus 1 in bovine semen. J Virol Methods. 2007 Sep;144(1–2):103–8.

7.	Boxus M, Letellier C, Kerkhofs P. Real Time RT-PCR for the detection and quantitation of bovine respiratory syncytial virus. J Virol Methods. 2005 May 1;125(2):125–30.

8.	Platts-Mills JA, Liu J, Gratz J, Mduma E, Amour C, Swai N, et al. Detection of Campylobacter in Stool and Determination of Significance by Culture, Enzyme Immunoassay, and PCR in Developing Countries. J Clin Microbiol. 2014 Apr;52(4):1074–80.

9.	Liu J, Gratz J, Amour C, Kibiki G, Becker S, Janaki L, et al. A laboratory-developed TaqMan Array Card for simultaneous detection of 19 enteropathogens. J Clin Microbiol. 2013 Feb;51(2):472–80.

10.	Tedim AP, Merino I, Ortega A, Domínguez-Gil M, Eiros JM, Bermejo-Martín JF. Quantification of bacterial DNA in blood using droplet digital PCR: a pilot study. Diagn Microbiol Infect Dis. 2024 Jan 1;108(1):116075.

11.	Yoshitomi KJ, Jinneman KC, Weagant SD. Optimization of a 3′-minor groove binder-DNA probe targeting the *uid*A gene for rapid identification of *Escherichia coli* O157:H7 using real-time PCR. Mol Cell Probes. 2003 Dec 1;17(6):275–80.

12.	McQuillan JS, Wilson MW. Recombinase polymerase amplification for fast, selective, DNA‐based detection of faecal indicator Escherichia coli. Lett Appl Microbiol. 2021 Apr 1;72(4):382–9.

13.	Verweij JJ, Blangé RA, Templeton K, Schinkel J, Brienen EAT, van Rooyen MAA, et al. Simultaneous Detection of Entamoeba histolytica, Giardia lamblia, and Cryptosporidium parvum in Fecal Samples by Using Multiplex Real-Time PCR. J Clin Microbiol. 2004 Mar;42(3):1220–3.

14.	Rudko SP, Ruecker NJ, Ashbolt NJ, Neumann NF, Hanington PC. Enterobius vermicularis as a Novel Surrogate for the Presence of Helminth Ova in Tertiary Wastewater Treatment Plants. Appl Environ Microbiol. 2017 May 17;83(11):e00547-17.

15.	Hidaka A, Hokyo T, Arikawa K, Fujihara S, Ogasawara J, Hase A, et al. Multiplex real-time PCR for exhaustive detection of diarrhoeagenic Escherichia coli. J Appl Microbiol. 2009 Feb;106(2):410–20.

16.	Costafreda MI, Bosch A, Pintó RM. Development, Evaluation, and Standardization of a Real-Time TaqMan Reverse Transcription-PCR Assay for Quantification of Hepatitis A Virus in Clinical and Shellfish Samples. Appl Environ Microbiol. 2006 Jun;72(6):3846–55.

17.	Cashdollar JL, Brinkman NE, Griffin SM, McMinn BR, Rhodes ER, Varughese EA, et al. Development and Evaluation of EPA Method 1615 for Detection of Enterovirus and Norovirus in Water. Appl Environ Microbiol. 2013 Jan;79(1):215–23.

18.	Boehm AB, Wadford DA, Hughes B, Duong D, Chen A, Padilla T, et al. Trends of Enterovirus D68 Concentrations in Wastewater, California, USA, February 2021–April 2023. Emerg Infect Dis. 2023 Nov;29(11):2362–5.

19.	Zhu K, Suttner B, Pickering A, Konstantinidis KT, Brown J. A novel droplet digital PCR human mtDNA assay for fecal source tracking. Water Res. 2020 Sep 15;183:116085.

20.	Green HC, Haugland RA, Varma M, Millen HT, Borchardt MA, Field KG, et al. Improved HF183 Quantitative Real-Time PCR Assay for Characterization of Human Fecal Pollution in Ambient Surface Water Samples. Appl Environ Microbiol. 2014 May;80(10):3086–94.

21.	Tsai YL, Tran B, Sangermano LR, Palmer CJ. Detection of poliovirus, hepatitis A virus, and rotavirus from sewage and ocean water by triplex reverse transcriptase PCR. Appl Environ Microbiol. 1994 Jul;60(7):2400–7.

22.	Zeng SQ, Halkosalo A, Salminen M, Szakal ED, Puustinen L, Vesikari T. One-step quantitative RT-PCR for the detection of rotavirus in acute gastroenteritis. J Virol Methods. 2008 Nov;153(2):238–40.

23.	Malorny B, Paccassoni E, Fach P, Bunge C, Martin A, Helmuth R. Diagnostic Real-Time PCR for Detection of Salmonella in Food. Appl Environ Microbiol. 2004 Dec;70(12):7046–52.

24.	Liu X, Yamamoto D, Saito M, Imagawa T, Ablola A, Tandoc AO, et al. Molecular detection and characterization of sapovirus in hospitalized children with acute gastroenteritis in the Philippines. J Clin Virol Off Publ Pan Am Soc Clin Virol. 2015 Jul;68:83–8.

25.	Lu X, Wang L, Sakthivel SK, Whitaker B, Murray J, Kamili S, et al. US CDC Real-Time Reverse Transcription PCR Panel for Detection of Severe Acute Respiratory Syndrome Coronavirus 2. Emerg Infect Dis. 2020 Aug;26(8):1654–65.

26.	Vu DT, Sethabutr O, Von Seidlein L, Tran VT, Do GC, Bui TC, et al. Detection of Shigella by a PCR assay targeting the ipaH gene suggests increased prevalence of shigellosis in Nha Trang, Vietnam. J Clin Microbiol. 2004 May;42(5):2031–5.
